# A qualitative investigation of views on practice in early support hubs from staff working in care and support roles

**DOI:** 10.1101/2025.10.22.25338534

**Authors:** Abbey Couchman, Rebecca Appleton, Kylee Trevillion, Hannah Kate Lewis, Connor Clarke, Jialin Yang, Phoebe Barnett, Sadiya Begum, Anam Bhutta, Julian Edbrooke-Childs, Eva Driskell, Jessica L. Griffiths, Isabel Hanson, Nima Cas Hunt, Emma Maynard, Lizzie Mitchell, Rob Saunders, Polly Waite, Brynmor Lloyd-Evans, Sonia Johnson

## Abstract

Early Support Hubs have recently become widespread in the UK and aim to provide community-based, easy access mental health support to young people aged 11-25, integrating a variety of forms of support. Evidence is needed on the role such services aim to fulfil in addressing young people’s mental health needs, perceived good practice in their operations and challenges encountered in achieving this. In order to understand this, we conducted individual interviews with 24 staff members from eight Hubs across England. We analysed data using codebook thematic analysis, identifying five themes: providing varied and holistic support; supported signposting and referrals; visibility and accessibility of hubs; important values embedded in the model, and remaining unmet needs. All participants felt that receiving care from Early Support Hubs could improve young people’s mental health, wellbeing and social circumstances. Important values guiding the practice of Hubs included being youth-centred, providing easily accessible and comfortable safe spaces, and employing staff whose approach is in keeping with Hubs’ values and ethos. Barriers to implementing the intended model included inadequate staffing levels, limited opening hours, and high staff turnover. Where they could not meet young people’s mental health needs, an intended role of the Hubs was to support them to find appropriate help: however, young people with significant mental health difficulties referred on by the Hubs sometimes did not meet high thresholds for receiving treatment from local mental health services. Research is needed to further understand the Hubs’ role in the system as a whole, their overall impact on addressing the rising burden of young people’s mental ill health, and how well-functioning local service systems that do not result in significant gaps in provision can be established.

## Introduction

There has been a global rise in mental health problems in young people (1–4) and three quarters of all lifetime mental health problems emerge before the age of 25 (5,6). Mental health problems during youth are associated with high recurrence rates (7,8) and poorer mental health, health related quality of life and life satisfaction in adulthood (9, 10). Despite the high prevalence and long-term impacts of mental illness, young people are the least likely age group to seek help (11–13) and international data suggests only around 20-30% of young people with a mental health problem receive an appropriate level of care to meet their needs (14–16).

Help-seeking during adolescence and early adulthood can be impeded by several factors. These include a lack of information about mental health and available services, perceived social stigma, not believing problems are severe enough for treatment and concerns around confidentiality and disclosing their problems to someone new (17–20). Even when help is sought, other systemic issues such as a lack of available services, long waiting lists, inaccessible locations and poor continuity of care further prevent or delay access to mental health care for young people (17). Poor continuity of care is often exacerbated by the structure of mental health services (21), with many children’s services stopping at 18, and a new referral required for adult services (22–25).

Integrated youth mental health services, a potential model of early intervention, have been developed internationally to increase access to mental health and wellbeing support for young people (26–28). They offer easy access, young person-centred, and responsive community-based care for young people, typically aged 11 to 25, who are experiencing the early stages of a range of mental health difficulties (29). Established examples of this model include Headspace in Australia (30–31), @ease in The Netherlands (32) and Jigsaw in Ireland (33). In England, comparable services are referred to as Early Support Hubs and are usually led by voluntary sector organisations, adopting a relatively non-clinical approach. They tend to lean more towards a non-clinical and community-oriented model and offer less structured therapy than the comparative international models listed above. Hubs can offer a variety of different types of support, including support for mental health and wellbeing, housing, financial advice, sexual health and careers advice (Youth Access) (34). There currently is not a clear model of what the primary aims are for these integrated youth mental health services and how these are to be achieved (34), despite a proposed national roll out of this model (35). Our aim in this study, together with a linked investigation of service users (36) and managers perspectives (37), is to understand the ways in which these services are aiming to help young people with their mental health and wellbeing, how far this can be achieved from stakeholder perspectives, and what impediments there are to achieving it.

Our main objectives are:

1. To explore staff views on the purpose of Early Support Hubs
2. To find out how staff perceive the benefits of this model of support for young people with diverse mental health, wellbeing and social needs
3. To identify challenges encountered in achieving the Hub model’s aims

## Methods

This research forms part of the larger NIHR Policy Research Unit in Mental

Health’s Early Support Hubs evaluation, commissioned by the Department of Health and Social Care (DHSC) in England (38) to inform national policy development. The write up of this study has been informed by principles of the COREQ checklist (39). Ethical approval was gained from the UCL Research Ethics Committee (27031/001).

### Setting

Individual qualitative interviews were conducted with staff working in a range of Early Support Hubs in England between June and August 2024. Participants were recruited from the 24 Early Support Hubs in England that received additional funding to develop their mental health support offer from the DHSC (38).

### Participants

Any staff whose role involved delivering direct care to young people were eligible for our study. To obtain a wide range of perspectives, purposive sampling (40) was used to ensure diversity in gender, ethnicity and role. We initially aimed to recruit 15-20 participants to capture a range of perspectives.

Information about the study, in the form of a poster and an invitation email, was sent to Hub managers to disseminate to their staff. Staff could either contact the researchers directly via email to express interest or have their contact details passed on by their managers. Researchers (AC, KT, RA) then provided potential participants with further information about the study via email, including a participant information sheet and consent form. Participants were encouraged to ask the researchers any questions about the study and written or verbal informed consent was obtained prior to each interview. After the initial phase of recruitment, we had interviewed 18 participants, but the sample was not ethnically diverse. We therefore extended the recruitment target to 25 participants to include the views of staff from ethnically minoritised backgrounds. No participants had any prior relationship with any member of the research team. Participants received a £20 shopping voucher to thank them for giving their time.

### Data collection

Interviews were conducted via Microsoft Teams by researchers AC, CC, and RA. Interviews followed a semi-structured topic guide (S1 Appendix). The topic guide was co-developed with members of the working group, including policymakers from the DHSC, academic researchers and two young people with lived experience, to ensure the questions were relevant to study aims and worded appropriately. Topics covered included: the type of care and support delivered by staff, how staff perceived the successes and challenges of their work, and what an ‘ideal’ early support hub model would look like. Interviews were recorded and transcribed verbatim using Microsoft Teams. Transcripts were then checked for accuracy by a member of the research team. After each interview, researchers made reflexive notes to consider initial thoughts and feelings from each interview during the analysis.

### Data analysis

Six researchers (AC, CC, KT, RA, HKL, JY), including a lived-experience researcher (HKL), undertook analysis of transcripts using NVivo 14. The first author (AC) analysed 10 transcripts, with the rest divided amongst the rest of the team. A codebook approach was adopted (41) to facilitate the application of inductive and deductive thematic analysis; the latter approach was used to generate pre-determined codes that aligned with the interview topic guide, and which enabled us to answer some key areas of interest to policymakers. A coding frame was used to record and chart the developing analysis, as well as guiding data coding for the multiple coders.

Initially, three researchers (AC, KT, RA) independently coded the same three transcripts before agreeing on a preliminary coding framework. This initial coding frame was then reviewed with the wider research team, and further iterations were made to the coding frame following these discussions. One researcher (AC) subsequently created a codebook which other researchers in the study team (AC, CC, JY, and HKL) then applied to the remaining interview transcripts. The approach to analysis was collaborative, with regular meetings held between the researchers coding the data to discuss arising themes and to agree on new codes to be incorporated into the coding frame. The process of reading, coding, discussion, and alteration to adapt the codebook continued between these researchers, with input from the whole team, until all data had been coded. Refining and naming the themes was the final stage of the analysis, which was led by AC and RA, with input from the wider study team.

### Reflexivity

The lead researcher, AC, is a female MSc student with a background in mental health support work, who conducted this research as part of her MSc and was supported by RA and KT. RA is an experienced female mixed methods researcher, with expertise in applied children and young people’s mental health research. KT is a Senior Lecturer and experienced female qualitative researcher. SJ is a female senior academic who is also a psychiatrist primarily working with young people with psychosis and bipolar disorder. CC is a male research assistant and HKL is a female lived experience research fellow. This project was informed by working closely with our team of Lived Experience Researchers, comprised of three young people with experience (LM, NCH and SBS) and a parent with experience of supporting their children with mental health difficulties (ED). The combination of academic, lived experience and clinical expertise in the team meant the analysis was informed by diverse perspectives.

## Results

Interviews were conducted with 24 staff from eight Early Support Hubs across seven different regions in England. The interviews lasted 43 minutes on average (range 25 to 57 minutes). Further sample characteristics are shown in Table 1.

**Table 1.**
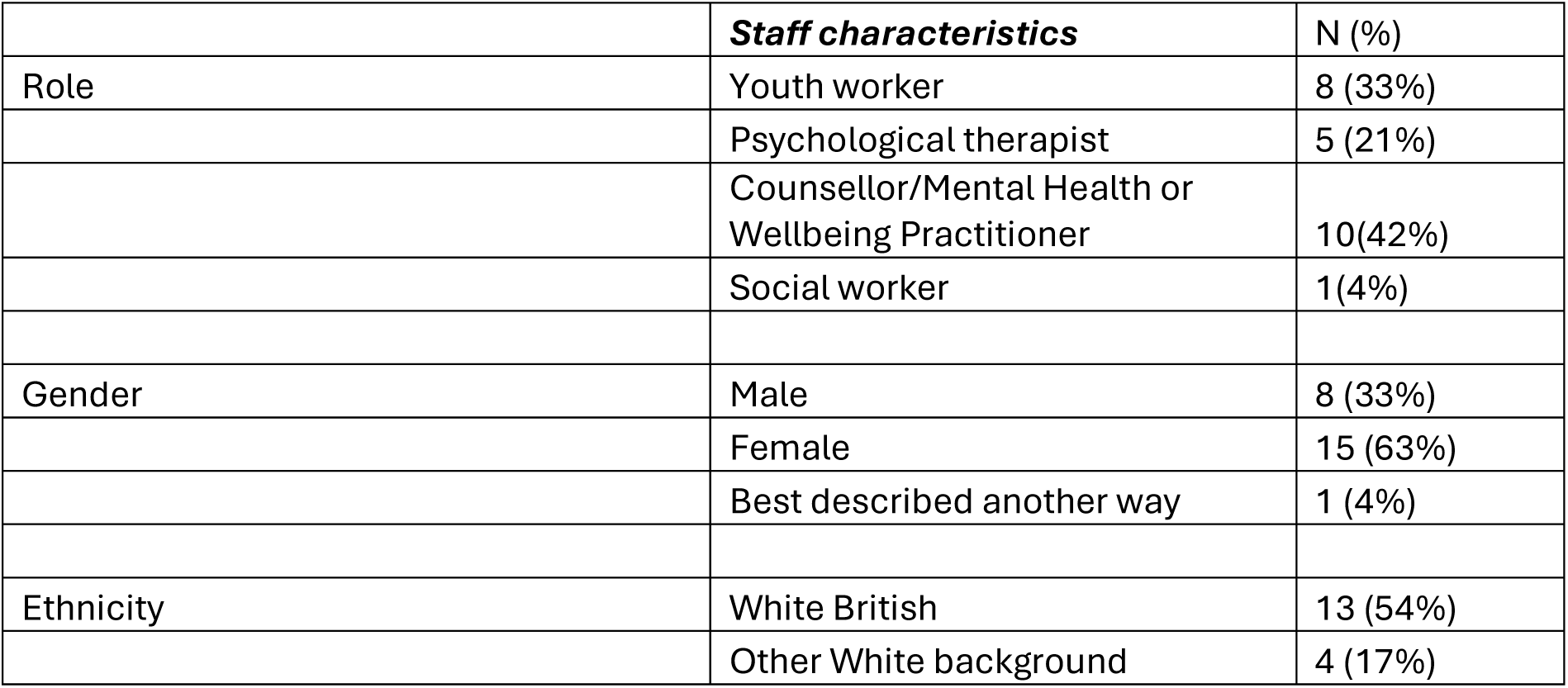
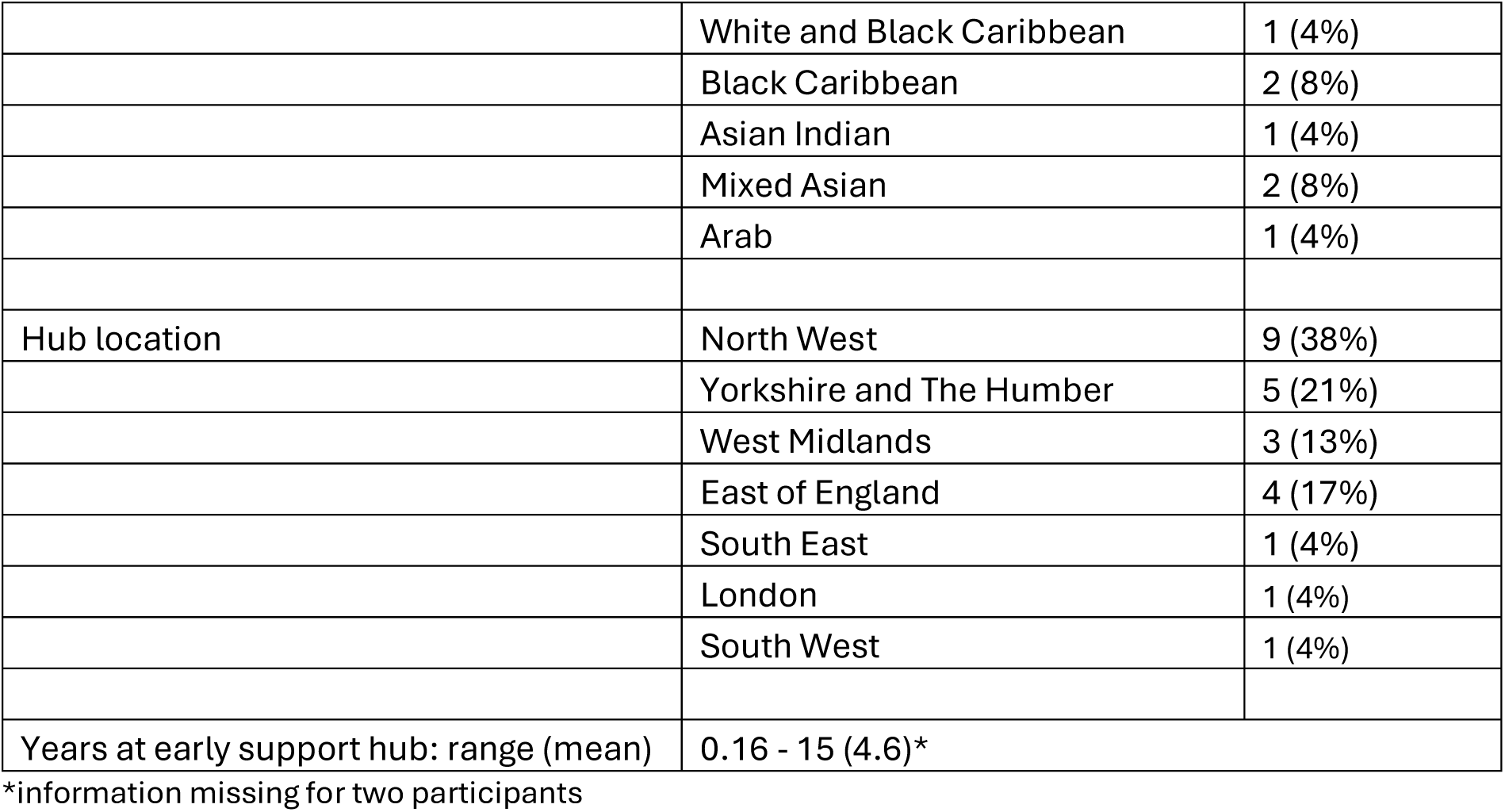
Professional and demographic characteristics of participants (N=24)

Five main themes were identified: providing varied and holistic support; supported signposting and referrals; visibility and accessibility of hubs; important values embedded in the model and remaining unmet needs. These themes, along with their subthemes, are summarised below, with illustrative participant quotes.

### Providing varied and holistic support

The types of care and support offered differed considerably between Hubs. Staff reported offering a wide range of interventions and activities for young people, with Hubs aiming to tailor their support to meet the needs of their local community. This was done by consulting young people about their needs and the kind of support they want, actively seeking their feedback, and remaining flexible and responsive in how support is provided.

> *“I think one of the other things as well, that’s a positive is we can be responsive, we’re more flexible. We don’t, we’re not trying to turn like a massive tanker in a bigger system. So we can be responsive to changes in the needs of the feedback or the presentations that we’re seeing within the community quite quickly.”* [participant 14, psychological therapist]

Some Early Support Hubs focused on low-intensity drop-in support, without the need for a pre-arranged appointment. Other services also provided more structured, longer forms of counselling or therapy (either as part of a group, or on a 1:1 basis), which were generally between six to twelve sessions in length, although one Hub reported offering 24 sessions. The types of therapy mentioned by staff varied across the different Hubs, and included solution-focused therapy, interpersonal psychotherapy, cognitive behaviour therapy (CBT) or therapy informed by CBT principles, or creative therapies including art, drama or music. In addition, some Hubs provided a space for young people to socialise, for instance, by having the drop-in area set up with snacks and games or running a youth club in the Hub. Activity-based support such as cooking, crafts or walking groups were also offered at some Hubs. Staff also described supporting young people with anxiety around school attendance, transitioning between primary and secondary school, bullying, and helping older attendees with social needs such as employability or finding housing.

Most Hubs provided drop-in support during weekday afternoons and evenings, with some also open on weekends. If a need for more intensive support was identified by staff during a drop-in session or triage appointment, Hubs would either refer young people to counselling/therapy internally, or signpost young people on to other relevant local services.

Most Hubs also provided specialist support for some specific needs, varying across Hubs. This included an integrated domestic abuse service, as well as groups for young carers, young parents, neurodiverse young people, young men, the LGBTQ+ community, refugees and asylum seekers and bereaved young people.

Overall, staff reported that the different types of care and support they provided allowed them to take a holistic approach. This holistic approach was seen to be a key function of Early Support Hubs, and something which sets them apart from other types of mental health services for children and young people:

> *“That’s kind of the function of the [Hub] really. We want to be like holistic support for young people and the Youth in Mind offer, which is like the new well-being offer is like the mental health side of that. But we already had projects for young people with additional needs, for socialising, projects for employability, projects for you know everything basically.” [participant 3, youth worker]*
>
> *“Ideally, Early Support Hubs are offering support around multiple issues. People usually come and our data would back this up with more than one thing that they want to help about. And I think that that’s very important that Early Support Hubs look at the whole picture of the person seeking help.” [participant 12, youth worker]*

### Supported signposting and referrals

Several participants highlighted that signposting and onward referral is a key aspect of the Hubs’ role and is achieved effectively when young people are supported through the process, emphasising that they *“won’t ever just signpost a young person and send them off and off they go. It will be a, you know, do you want to write an e-mail together, or should we book you an appointment and then I’ll check in with them afterwards and see how it went, or do you want to let me know?” [participant 2, youth worker/team lead].* This approach was felt to be beneficial in helping young people access the appropriate type of support to meet their needs:

> *“I think that we also are really useful in helping young people navigate more complex services and systems as well as helping young people really like unpick, what kind of support they want and need and then helping them get that.” [participant 12, youth worker/team lead]*

The effectiveness of signposting and referrals was enhanced when Hubs had established relationships with other services. For instance, one participant attributed their confidence in handling risk to the clinical supervision they received from CAMHS practitioners.

> *“I also have the flexibility that if I feel like I’m not equipped to help a young person, then I am just, like I can research. I can talk to people here, I can talk to people at CAMHS. We have supervision from CAMHS. So, if I’m ever not sure about working with a young person or I’m not sure about the risk or what to do with them, then I can organise a Teams call with somebody from CAMHS and talk it through from their perspective as well.” [participant 3, youth worker]*

Furthermore, participants described how the Hubs’ relationships with other services could allow a more holistic approach than the Hub could offer on its own:

> *“We will quite often take that multidisciplinary team approach to give that young person wraparound holistic support, so we will work with social prescribers, we will work with teachers, we will work with social workers, family youth workers, to start taking a collaborative approach.” [participant 8, youth worker]*

Participants also described how they would like to strengthen their connections with other services, if these connections were not already in place:

> *“I would really like to strengthen our relationship with other teams like the housing advisory team and the social care team to make sure that we’re doing the best we can and working as quickly as we can as well.” [participant 12, youth worker]*

### Visibility and accessibility of Hubs

Staff recognised the need for young people to be aware of Early Support Hubs and the support they offer. Many participants saw simple *“word of mouth”* [participant 2, youth worker] within the community as important in achieving this. Effective outreach work was also an important factor in visibility; some participants cited this as a strength of their model, whereas others felt they needed to do this more extensively:

> *“I feel like we offer such an amazing service that the whole city should know about it, and as soon as a parent knows or recognises that their child has got kind of any anxiety issues or young adults have noticed that they need require support or something, [Hub] …should be on everybody’s mind.” [participant 4, youth worker]*

The accessibility of support offered by Hubs was identified as important by several participants, both in terms of support being easily accessible without the need for a referral and by being in physically accessible buildings, based in easy-to-reach central locations. Several participants commented that physical accessibility, affected by the building (e.g. not having a lift) and location of the Hub, was an important consideration. However, this was an area that needed improvement at many Hubs.

> *“I recognise that although our Early Support Hubs are accessible, they’re not accessible to everyone, not everyone has the money to be able to just get the bus.” [participant 19, Mental health or Wellbeing practitioner]*

The need for services to feel approachable was widely reported, with a few participants indicating that running a youth club as part of the Hub was a good way of making mental health support visible and approachable for young people:

> *“It means that young people can go to a central place to access lots of different types of support because for some young people just getting through the door of somewhere new is a barrier due to anxiety. So the fact that they’ve already sort of built a relationship with somebody here and then they can tap them into other services is amazing.” [participant 8, youth worker]*

By providing easily accessible and approachable, low-intensity care, Early Support Hubs are seen as filling an important gap that results from the high threshold for specialist services:

> *“I feel like [the] Early Hub is like a more accessible CAMHS because the way I used to work in CAMHS is only by appointment. You can’t just walk in, it’s just like the GP, more clinical. But to me I feel [the] Early Support Hub here, you get similar kind of therapy but it’s, less threatening and it’s more approachable.”* [participant 23, psychological therapist]

### Important values embedded in the model of an Early Support Hub

#### Young person-centred

All participants stressed that it is important for Early Support Hubs to be young person-centred, both in terms of the individual care received and the design of the service. Participants regularly used the term ‘youth-led’ to describe their Hub. There was an emphasis on providing stigma-free and non-judgemental support for young people, and making sure they are “*feeling truly heard*” [participant 17, social worker]

Participants contrasted the Hubs’ youth-centred approach - *“it’s really not tokenistic” [participant 12, youth worker]* - to other services which they described as strict, clinical, inflexible, and not youth-friendly. Some Hubs ran participation groups to facilitate young people’s input to Hub design and delivery, for example, through interviewing prospective staff, and helping to design new activities or types of support offered. One participant also described a new group therapy programme which will be delivered by young people and supervised by staff.

> *“Rather than it just being us kind of putting this programme together and hoping it meets the needs, it’s actually about hearing the young people’s voices and what they need. And constantly doing consultations with young people”. [participant 8, youth worker]*

Many participants viewed young people, rather than staff, as the experts on their own needs. Accordingly, when asked about how they would improve the Hubs, many participants emphasised the need for ideas to come from the young people.

> *“I’d like to do a real big participation piece in the community with the young people to see what they want. And see what works for them and what has worked, what hasn’t worked… they would be part of that design”. [participant 14, psychological therapist]*

However, one participant raised some limitations of the youth-centred model, as it can result in young people focusing on areas that are within their comfort zone, avoiding others that may be important but more uncomfortable to address:

> *“And I think it’s also that tricky thing, like we might do some work with the young person because we’re so young person centred, we have to go with what they want and their priorities. But sometimes you’re working with someone and you kind of think like, OK, you want to come in here and talk about your friendship issues but what about the fact that you haven’t been to school for three weeks, but they don’t want to talk about that? So sometimes I feel like because we’re so young person-centred, we might not kind of maybe step up and challenge them in a way that maybe they need to be challenged. So to kind of be like rather than talking about what’s going on Snapchat, should we talk about how we could get you into school?” [participant 1, youth worker]*

Flexibility was also seen by some staff as a crucial element of a youth-led approach, allowing the approach used, the number of sessions delivered, or types of support offered in combination to be tailored to each individual:

> *“…the premise of [Hub] is we’re flexible, we meet the needs and it’s, it’s what we call child, person centred, client-led and it’s meeting their needs. So we adapt and we’re collaborative with the young person.” [participant 14, psychological therapist*

Staff described having a flexible way of working with young people, allowing them to engage with support on their own terms. For example, Hubs would continue to offer support to young people if they missed several sessions, which was contrasted with other statutory services. Some staff stated that support should only be provided if the young person actively sought it and was ready to engage, and not at the request of their parents, schools, or other services.

This approach was sometimes acknowledged as challenging for parents who may be concerned about their child’s mental health:

> *“…there are obviously times where parents bring their children into us and the children don’t want to see us. And that’s really difficult because you can see how much the parent is struggling. But within our guidelines we have, we can only support a young person that wants the support from us.” [participant 4, youth worker*

### Promoting independence

Participants saw their roles as helping young people to fulfil their potential and to become more independent. One staff member described the purpose of the Hub’s drop-in advice service as being to help young people navigate new situations and challenges, such as applying for jobs or getting access to social housing:

> *“I would say the purpose is to like help young people transition to adulthood and answer those questions that like you just don’t know how to do those things because you’ve never done it before.”* [participant 1, youth worker]

Encouraging independence and the development of new skills was seen as important: *“giving them tools and really empowering them really to become their own, if you like, therapist.” [participant 7, counsellor]*.

> *“I think that is, and to be able to thrive and reach that potential and to be able to learn coping mechanisms to be able to manage the mental health. And again, for that to be integrated outside of this space so that they don’t need us anymore, that’s kind of our job is to go, you don’t need it anymore, you’ve got this.”* [participant 8, youth worker]

### Anyone, any way

Easy access was also seen as central by many, with no one turned away because they did not meet criteria, and help made available immediately:

> *“No problems too big or too small for people to bring to mental health advice.” [participant 1, youth worker]*

As well as being available to everyone, participants described how the drop-in provisions allow young people to access help immediately, as and when it is needed, and to easily obtain follow-up support via repeat attendance.

> *“They might always have a bad day or a bad week, or a bad month but because we’re an open service… they can always come back to us. We’ve got an open door and they know that.” [participant 8, youth worker]*

This was contrasted with long waits for other mental health services: *“I’ve certainly worked with a lot of young people that have been on for example like NHS [National Health Service] wait lists or wait list for counselling with other third sector organisations. And it’s kind of like, I guess you get a letter that you’re on the wait list and then you just wait for however many months or even years until that happens, whereas at an Early Support Hub there are like multiple points of entry you can come back whenever you want…”* [participant 12, youth worker]

### Providing a comfortable and safe space

Staff also saw a welcoming and comfortable environment as being important, contrasting this with more clinical atmospheres in traditional services. Involving young people in decorating Hubs was seen as a good way to achieve this:

> *“They are welcoming warm places that you know are comfortable and like homely, almost like they feel like they’ve been designed with young people in mind. It’s not like a doctor’s office or something.” [participant 12, youth worker*

In addition to the physical space, participants discussed the importance of creating a psychologically safe space. The majority of participants felt they did this successfully, partly due to building strong and trusting relationships with the young people.

> *“I feel really confident that young people who access us, they come here, and they feel welcome, and they feel heard. And I think those are the two most important things in a help seeking experience.” [participant 12, youth worker]*

Some participants felt that the creation of a psychologically safe space was also facilitated by their confidentiality from schools, parents, and carers. Participants from one Early Support Hub were especially averse to involving parents or carers in the support they provided as their *“focus is on what that young person is sharing” [participant 2, youth worker]* and would only involve parents at a young person’s request. However, Hubs varied – for others, working with parents and carers (either alongside the young person, or separately) was a key part of their model.

> *“We can fully do the same session we would do with young people to parents in terms of giving them the skills or strategies to kind of help improve the situation. It may be that we just kind of come and have a conversation around what’s been going on and find them the best support as we’d have more kind of knowledge around it or more kind of options. Sometimes it can just be some reassurance towards parents, or even giving them a bit of space while a young person comes in and sits on a beanbag or plays a board game. Or even, as I said, just coming in and being able to offload.” [participant 10, Mental health or Wellbeing practitioner]*

### Staff teams with the right values, skills, and support

Many participants described the importance of staff having the right qualities and values which are compatible with the ethos of the Hub, such as having a caring nature, and having prior experience working with children and young people. This was seen as important when hiring staff, rather than prioritising their qualifications. This was felt to improve the quality of care provided to young people.

> *“It’s about kind of having staff who might have, you know, a range of staff that have got different skills, different practises and that have got a range of different life experiences themselves as well. So, the young people can see themselves reflected in the staff force.” [participant 13*, psychological therapist]

The diversity of staff in terms of expertise and experience facilitated working collaboratively as a team, as staff could draw on each other’s strengths.

Participants described the Hubs workplace culture as supportive and friendly.

This was seen as especially important in the context of working in a fast-paced, emotionally demanding environment. The depth and breadth of training provided by the hubs, informal support from colleagues, and an *“open door policy”* [participant 13, psychological therapist] from management facilitated the supportive environment.

Team meetings and peer support groups were also frequently identified as sources of support for staff. Some participants also highlighted the value of external supervision (e.g. from CAMHS professionals) to learn from other perspectives.

> *“The access to kind of training and supervision and I think is really good as well. And the informal support that you get at [the Hub] as well. It’s a nice organisation. People get on with each other. Which I think I know that sounds like a small thing, but actually it’s a massive thing. Like it’s a good team with people that hold the same kind of values. So, you can kind of turn around to a colleague for informal support.” [participant 13, psychological therapist]*

### Remaining unmet needs

#### Cannot, or Shall Not, Fill the Gap

Due to the easy-access nature of Hubs, participants sometimes saw young people with a high level of mental health need such as young people who are severely distressed, or experience suicidal ideation, or presenting with symptoms of eating disorders, OCD, or psychosis, who may require more specialist treatments. Many participants felt they could successfully support these young people by providing supported signposting and referring people to specialist support from other services. Staff also spoke about having to call crisis or emergency services for young people who needed urgent support.

Some participants, however, felt they were unable to offer appropriate support within the Hub, particularly for young people with a high level of social need:

> *“So sometimes it feels like actually we are becoming obviously not social workers, because that’s not our job, but it feels like we are the only ones with kind of eyes on a very vulnerable young person and having to kind of connect them into different agencies and things like that where I think we probably experience young people with a lot higher needs than expected, so that can be challenging sometimes.”* [participant 8, youth worker]

There were different views around whether Early Support Hubs should be able to offer more intensive support to these young people, or whether this is beyond their role. A few participants described a gap emerging in which CAMHS are only able to support those with the highest level of need, and Hubs are designed to support a low level of need, leaving young people in the middle unable to access the appropriate support. For example, one participant described receiving referrals from early intervention in psychosis services due to a young person not meeting the service’s criteria, but feeling unable to give them adequate support themselves, leaving the young person going “*round and round in circles, and in and out of services.*” [participant 13, psychological therapist]. Therefore, some participants suggested that it would be useful to offer a higher level of support at the Hub and *“challenge the idea that Early Support Hubs are just for early intervention.” [participant 12, youth worker]*

Some identified a need for additional training, for example, in eating disorders and OCD, if Hubs are to provide this more specialist support.

In contrast, other participants felt it would be more useful to increase the capacity of CAMHS and adult community services.

> *“Early Support Hubs are meant to prevent young people from getting into crisis, however, we are noticing a rise in crisis presentations within our early support hubs, so I suppose with that in mind, I would think that there’s need for more crisis services, more funding for community mental health, more funding for youth services, and more funding for NHS.” [participant 19, Mental health or Wellbeing practitioner]*

A few participants explicitly recognised the importance of ensuring the Hubs’ model fits into the local context of care. This allows the Hubs to fill local gaps in service provision, work efficiently with other services, have clear pathways for young people, and ensure equitable distribution of workload.

> *“I think there needs to be the understanding of where, how the Hub fits into the community and into the area in terms of other services… and looking at actually, okay, so what, what can you offer, what do you offer and then what do we offer and how it links and it works together. So, for me that’s kind of like the umbrella thing first.” [participant 14, psychological therapist]*

### Not Enough

All participants identified issues of capacity created by inadequate or unstable funding. Limited funding resulted in not having enough staff and reduced opening hours (especially at weekends). Participants felt like they could not *“see everybody that feels they need to be seen or wants to be seen.” [participant 2, youth worker].* Low staffing also meant that participants had to see young people back-to-back which was experienced as stressful. In addition, one participant described how they would “*end up doing things that are not within [their] job role”,* such as substantial safeguarding and inter-agency working *“but do them anyway because there’s no funding for someone else.”* [*participant 8, youth worker]*

This further increased the strain on staff; the limited and turbulent funding for Hubs also affected staff retention, with staff citing worries around job security and the financial implications of this. Concerns were also expressed about how the consequent high staff turnover and reliance on temporary staff disrupts the Hubs’ ability to maintain a consistent approach and hinders their long-term progress.

> *“It’s my biggest frustration is that stability that we that we need because everything we offer here is badly needed. You know, every service that we provide in this building is something that if we didn’t do it, our town would be the worst for it.” [participant 9, youth worker]*
>
> *“A service from beginning to end took at least three years to bed in, settle in. And if you’re doing 3-year funding roll on, roll off, it’s, it’s unsafe. It’s you know, it’s quite disillusioning for staff as well.” [participant 18, psychological therapist]*

Limited capacity created unique issues in drop-in services, as most Hubs ran the drop-in sessions with a first come, first served policy. This meant that there is a risk that not all young people who attend a drop-in session will be seen if the Hub is at capacity.

Similarly, participants from Hubs that offered psychological therapies and counselling reported longer waiting lists for these services ranging between 3-9 months which limited their accessibility, or the number of sessions offered.

> *“I think the waiting lists at our service are service are sort of really big. And we’ve got a lot of young people that need support in the city and consequently sometimes it feels as though you might not be able to work with the young person for as long as you might want to or might feel is sort of going to be the most helpful for them.”* [participant 13, psychological therapist]

One participant explained that, to reduce waiting lists, young people were assigned to the first available practitioner. However, this created issues as the young person and the practitioner (or the therapeutic approaches they were trained in) were not always the best match. Staff felt that that extra funding could reduce the time young people had to wait and help to achieve their aim of providing early intervention.

Similarly, a staff member from one Early Support Hub described how their service delivered a set model with a limited number of sessions, which they felt was not effective for some young people who required longer term support due to the symptoms they were presenting with. Staff reported frustrations with the lack of flexibility of this approach:

> *“It’s still an eight-session model, and yet sometimes we’re getting clients through with a lot of complexity. So, it’s kind of how do you meet the needs of those clients within the model of our service?… It kind of feels sometimes like we’re putting plasters on gunshot wounds.” [participant 13, psychological therapist]*

Some participants wished for extra funds to be able to provide additional comforts such as better snacks, games consoles, and comfortable furniture. At some Hubs, more basic fit-for-purpose facilities were needed, such as a dedicated waiting rooms, private room availability, sound proofing, and lifts, to ensure all areas were physically accessible for people using mobility aids, which participants felt all had a major impact on the quality of care.

> *“Sometimes we really struggle to get rooms… And then you’ve kind of got to either, you know, call the client back and try and reschedule or, which is obviously impacting on their needs.” [participant 13, psychological therapist]*

A lack of funding often adversely affected Hubs’ ability to support young people with more specialist needs. Predominant concerns were around supporting neurodiverse young people, young people with additional educational needs, or young people who may need help from a translator or sign language interpreter to access support. Participants had ambitions to run specific therapy groups, make changes to the physical environment, and conduct outreach to better support minoritised groups, but these often could not be implemented due to a lack of resources.

> *“It’s too busy for them. It’s too over stimulating. It’s too overwhelming. So, to be able to have a smaller support group for SEND young people I think would be absolutely incredible.” [participant 8, youth worker]*
>
> *“We previously did workshops where we collaborated with a project that worked with like refugees and stuff… those sets of workshops are over because the contract finished.” [participant 6, Mental health or Wellbeing practitioner]*

A need for more outreach work to meet the needs of minoritised groups in local communities was also raised by participants, including young people from Muslim or Traveller communities. One staff member suggested that forming closer links with churches and faith groups in local communities could improve access to their service for minoritised children and young people.

> *“I think reaching out to them in their communities. And being able to do some psychoeducation about the importance of accessing support because if we break it down and go to the starting point of accessing support, certain communities can struggle with accessing support, that asking for support can be really challenging for many different reasons. It could be for reasons that they have had negative experiences with professionals, and they felt unheard. Therefore they kind of isolate themselves and choose not to seek support when needed.”* [participant 24, Counsellor & Psychological therapist]

## Discussion

We found that staff in our study felt that the current UK model of Early Support Hubs makes a valuable and distinctive contribution to mental health support for young people in several ways. Participants identified several key factors that contributed to the success of an Early Support Hub and distinguished them from other mental health services, such as their youth-centred and holistic approach, accessibility (through self-referrals, drop-ins, having no minimum thresholds for access), flexibility, non-clinical space, and their diverse and compassionate workforce.

However, several tensions were also identified in the delivery of early support hubs. Staff described the challenge of providing easy-access, early intervention support to all young people whilst also ensuring the needs of those with higher levels of, or more complex, mental health difficulties are met, in the Hubs or elsewhere. There were also differing opinions on how maintaining a psychologically safe and confidential space for young people should be balanced with also involving and supporting parents and carers. Although the open-access policy was viewed as a key strength of the Hubs, staff reported ongoing difficulties in raising awareness of Hubs and effectively engaging the whole local community. Additionally, the short-term nature of Hub funding in this particular national programme and challenges in recruiting and retaining skilled staff who embody the necessary qualities and values impacted the Hubs’ capacity and long-term sustainability.

These findings from the current study mirror other qualitative research with staff and service users from international models of integrated youth services, such as on the importance of a youth-centred approach (42–44), a welcoming, non-clinical physical environment (45) and effective partnerships with other services (46). Staff working at Headspace in Australia have also reported tensions regarding their ability to support young people with moderate to severe mental health needs, a need for more specialist training, and unresolved questions as to whether that should be part of their role in an early intervention service (43). A need to better promote their services and outreach work for specific groups (e.g. young men) has also been reported elsewhere (43, 46), underscoring the need for youth early intervention services to ensure they are accessible and visible to all young people within their communities.

The key values identified by Hub staff align with young people’s preferences for mental health services as reported in wider literature (47), such as creating a welcoming environment and ensuring they feel heard and valued. These values also address several barriers to accessing support that young people have identified (17), including the need for a non-medicalised approach, practical support, and immediate access. Specialist mental health services in England have been described by young people, parents and clinicians as having disadvantages including inflexibility, poor processes for transition from children’s to adults’ services and lack of accessibility or early responsiveness: from the perspective of our study participants, the Hubs have potential to address these difficulties (23–25).

However, staff experiences suggested that Early Support Hubs currently do not fully meet the full range of preferences expressed by young people in previous studies. For instance, young people prefer to see the same member of staff (47), which is not facilitated by the drop-in model, or high levels of staff turnover, caused by short-term funding. Some Hubs also appeared limited in their capacity to publicise their service and conduct outreach work in their local communities, so that young people may not in practice know where to access support without the need for a referral (17). As not all Hubs were able to offer effective outreach work, there is a risk of widening existing inequalities in who is able to access early mental health support.

Although participants identified important values and aims for of the Early Support Hub model, these could not always be implemented in practice. Short-term funding underlined many of the Hubs’ limitations, such as capacity, retaining staff, being able to meet the needs of specific groups, and having a fit-for-purpose space. As short-term funding was cited as a major barrier to effective implementation of this model by staff in this study, there is a need to consider secure, long-term funding in the future. The NHS Long Term Plan (48) acknowledges the need to fund voluntary sector health services effectively to address these constraints.

Beyond funding issues, the Early Support Hub model had other limitations, such as their unclear role in supporting young people with a higher level of long-term need or in crisis. Hubs typically include staff from a range of backgrounds, and many will not be trained to work with young people with more severe mental health problems or where specialist skills are needed in areas such as psychosis or bipolar disorder. There is a need for clarity on whether Hubs should increase their offering to support young people with a higher level of need, or if other services fill this gap.

Participants expressed concerns that some young people may be unable to access appropriate care if support from the Early Support Hub does not meet their needs, yet they do not meet the thresholds for other NHS services. The gap in service provision has been well documented (49); for example, in 2022-23, 372,800 children and young people referred to CAMHS were not accepted for treatment (50). As a result, some staff felt that they had to provide this support within the hub. This sometimes meant performing tasks they felt were outside of their job role, which could be challenging. Some staff felt that the remit of early support hubs has been stretched too far, potentially losing its focus on early intervention.

### Strengths and Limitations

The study benefited from collaboration with lived experience researchers, clinicians and academic researchers with various perspectives from the beginning of the research process. In particular, the collaborative approach taken to data analysis which combined experiential, clinical and academic expertise led to enhanced data interpretation, as it allowed us to draw on varied experiences. The use of purposive sampling also resulted in diversity across participants in terms of their roles, previous experience, gender, and ethnicity, and we interviewed staff across several different Hubs.

Limitations include a potential implicit pressure on staff to report positive views of the model, especially given the insecurities of funding. Some staff also participated within their Hub’s office spaces, potentially restricting discussion about the model’s limitations further. The varied nature of support provided by Hubs makes it hard to articulate their exact model and intended pathways to providing effective mental health care, and we do not know whether our findings apply to all Hubs across the country.

This may be exacerbated by the fact that we only interviewed staff from eight of the 24 Hubs receiving additional funding from the DHSC and that other Hubs are not taking part in this evaluation, meaning our findings are unlikely to reflect the full range of views of Hub staff across the country. Hubs also offer a variety of different types of support to a wide age group of 11-25 years. However, staff were not always precise about the age range they were referring to in their responses, meaning we do not know if all findings apply across this age range.

### Implications for Practice and Research

The results of this study indicate that staff are able to identify clear ways in which the Early Support Hub model has potential to make a distinctive contribution within the overall landscape of mental health support for young people in England. The findings highlight large variations in the types of support offered by different Hubs, suggesting a need to distinguish different Hub models and, ultimately, define core components of a successful model including the required workforce to provide this support. Future research should explore the effectiveness of different types of support offered by Hubs in improving young people’s wellbeing and mental health. Young people who meet criteria for a variety of mental health diagnoses, including depression and anxiety disorders, are likely to be accessing the Hubs. However, Hubs may not offer standard treatments for depression and anxiety, as recommended in NICE guidelines (51–53), and their relatively limited range of clinical interventions may differentiate them from early intervention models described in the literature. Therefore, it is important to determine what types of support are being delivered and whether outcomes are at least as good as for other models of treatment for these conditions, in addition to what types of support work best for which young people.

Secondly, there is a need to consider the whole local service system and ensure there are care pathways available for all levels of need and acuity. This is important to enable all young people to access early support for mental health problems, particularly those who are experiencing difficulties that the Hubs are not able to support with but who fall below the threshold for CAMHS support. Effective partnership working is important to enable timely referrals between services, with clear guidelines for when an Early Support Hub will refer a young person on to a higher level of support, ensuring that young people do not fall through gaps between different services.

Adequate funding for surrounding services will allow hubs to focus on prevention and early intervention (32) and conceivably reduce the impact of mental illness long-term (54–56). This understanding of how services should fit together could be achieved through further qualitative research that explores the views of staff from surrounding services, such as CAMHS and other local charities.

Finally, in the context of a programme receiving short-term funding from the government to enhance its mental health provision, staff spoke of the importance of adequate and stable funding for Early Support Hubs if the intended aims are to be achieved. Sufficient funding would also enable Hubs to employ a permanent and skilled workforce, provide stable support without interruptions from funding cuts, increase their opening hours and reduce waiting lists for structured support. It would also allow them to conduct more outreach work, especially with minoritised groups, and have the flexibility to be responsive to the needs of young people in their local area (such as by running specific support groups), and to maintain high quality and physically accessible premises with sufficient space to deliver interventions.

## Conclusion

Early Support Hubs are a potentially promising model of providing various types of mental health and social support to young people in their local communities.

Participants spoke of the role of Hubs in filling gaps in local service provision, although it is still unclear how Hubs can best fit in with other mental health services and meet the needs of young people with more severe symptoms. Research is needed to further understand the Hubs’ role in local service pathways and their overall impact on addressing the rising burden of young people’s mental ill health.

### Lived Experience Commentary

#### Written by a family carer and a member of our NIHR Mental Health Policy Research Unit Working Group - Eva & Anam

We are well aware of the gaps in services and how Children and Young People (CYP) and their families and carers are left to struggle in isolation with sometimes frightening and dangerous mental health problems. This study shows how widely the services vary and highlights the need for best practice. The principles described by some of the staff interviewed are positive - holistic, youth-centred with immediate and open access.

#### Consistency

Consistency is crucial to meet the varied needs of CYP from all backgrounds and circumstances. Staff retention is critical - regularly changing key workers and having to rebuild relationships and repeat information is wearing and distressing.

#### Inclusion

Mental health is largely misunderstood and stigmatised in many cultures. Those from BAME backgrounds are more likely to struggle with their mental health, yet fewer people from these communities access support services. Many CYP may be hesitant to engage with Hubs - better support for parents and carers to view the Hubs in a positive light may lead to conversations of encouragement to their children to attend the Hubs. It is possible to include and educate the wider support network without compromising the safety and confidentiality of the CYP. The Triangle of Care model (service user, professional, carer) used across many adult mental health trusts ensures that all involved are heard, understood and work together. The needs of those in more rural locations must be considered and met e.g. accessing all services and support groups online or by phone.

#### Unmet needs

Staff need more training around how to appropriately respond to young people with more complex needs, as well as mental health struggles that are not well understood. For example, some important difficulties mentioned in the paper that staff reported an increase in presentations of and wanted more specialist training in were OCD, eating disorders and psychosis. Staff would benefit from receiving additional training to help them become aware of the different presentations of such disorders. Many of these disorders can co-occur and overlap with symptoms of other mental health disorders, making their presentations complex and requiring tailored treatments. Having trained staff who are already aware of the unspoken struggles within society and how to deal with these symptoms can make a real difference in making a young person feel safe, heard, and understood.

Finally, these Hubs should ease the transition at 18 between CAMHS and adult services, but are possibly pushing the problem down the line and consideration is needed for the transition at 25.

## Data Availability

The data that support the findings of this study are available on request from the corresponding author. The data are not publicly available due to privacy or ethical restrictions.

## Appendix

## S1 Appendix. Interview Topic Guide

### PART 1: CURRENT PROFESSIONAL ROLE AND EXPERIENCE

1. Could you please describe your current role in the early support hub and years of experience working at the early support hub and more broadly in care and support for young people? *Potential prompts to include:*

- Professional qualifications in health and social care

### PART 2: DESCRIBING EARLY SUPPORT HUBS

2. How would you describe what an early support hub and its purpose is to someone who doesn’t know about them? *Potential prompts to include:*

- Who is and isn’t the service intended for?
- What is the purpose of the hub/the intended model?
- What are the important characteristics of an early support hub?
- What types of care and support does the service provide for young people?

3. What are the needs of the young people who access the early support hub?
4. For those who have worked in any other mental health services for young people, how are early support hubs different to these?

### PART 3: EXPERIENCE OF WORKING IN EARLY SUPPORT HUBS

5. How successful do you think the early support hub is in working with young people with mental health difficulties? What helps or hinders this? *Potential prompts to include:*

- What works well for the young people? Which young people benefit?
- What doesn’t work well for the young people? Which young people don’t benefit?
- What works well for you and your role?
- What doesn’t work well for you and your role?
- What impact does the early support hub have?
6. [If appropriate] In what ways is the early support hub better or worse than other mental health services you have worked in?
7. Have you received any training whilst working at the early support hub? *Potential prompts to include:*

- What training was helpful or not helpful/relevant?
- Is there any other support that you would like to receive to improve the care and support you provide for young people?

### PART 4: FUTURE RECOMMENDATIONS FOR EARLY SUPPORT HUBS

8. Do you feel like you have an active role in service improvement? *Potential prompts to include:*

- Improvements regarding service delivery
- Improvements regarding accessibility
- Improvements regarding types of support
9. What improvements would you make to the early support hub, assuming resources are available? *Potential prompts to include:*

- What could be improved to make access to the early support hub easier for young people?
- Do the young people using hubs have any needs, which the hubs don’t help with, could anything be done to improve this, and how?
- What would you like to improve for people who work at the early support hub in care and support roles?
- Is there anything you have seen in other mental health services that would be useful to provide in your hub?

10. In an ideal world, what do you think an excellent service delivering early support for young people in mental distress in the community should look like? *Potential prompts to include:*

- Type of care and support offered (e.g. mental health care, physical health care, practical support) and any resources provided
- Accessibility and opening hours
- Who would work there?
- What kind of environment would it be? (e.g. regarding waiting room, the support rooms, the location)
- What impact would the early support hub have?

### PART 5: CONCLUDING QUESTION

Is there anything you would like to add that we have not yet spoken about today?

## S2 Appendix COREQ (COnsolidated criteria for REporting Qualitative research) checklist

Developed from: Tong A, Sainsbury P, Craig J. Consolidated criteria for reporting qualitative research (COREQ): A 32-item checklist for interviews and focus groups. International Journal for Quality in Health Care. 2007 Dec;19(6):349–57.

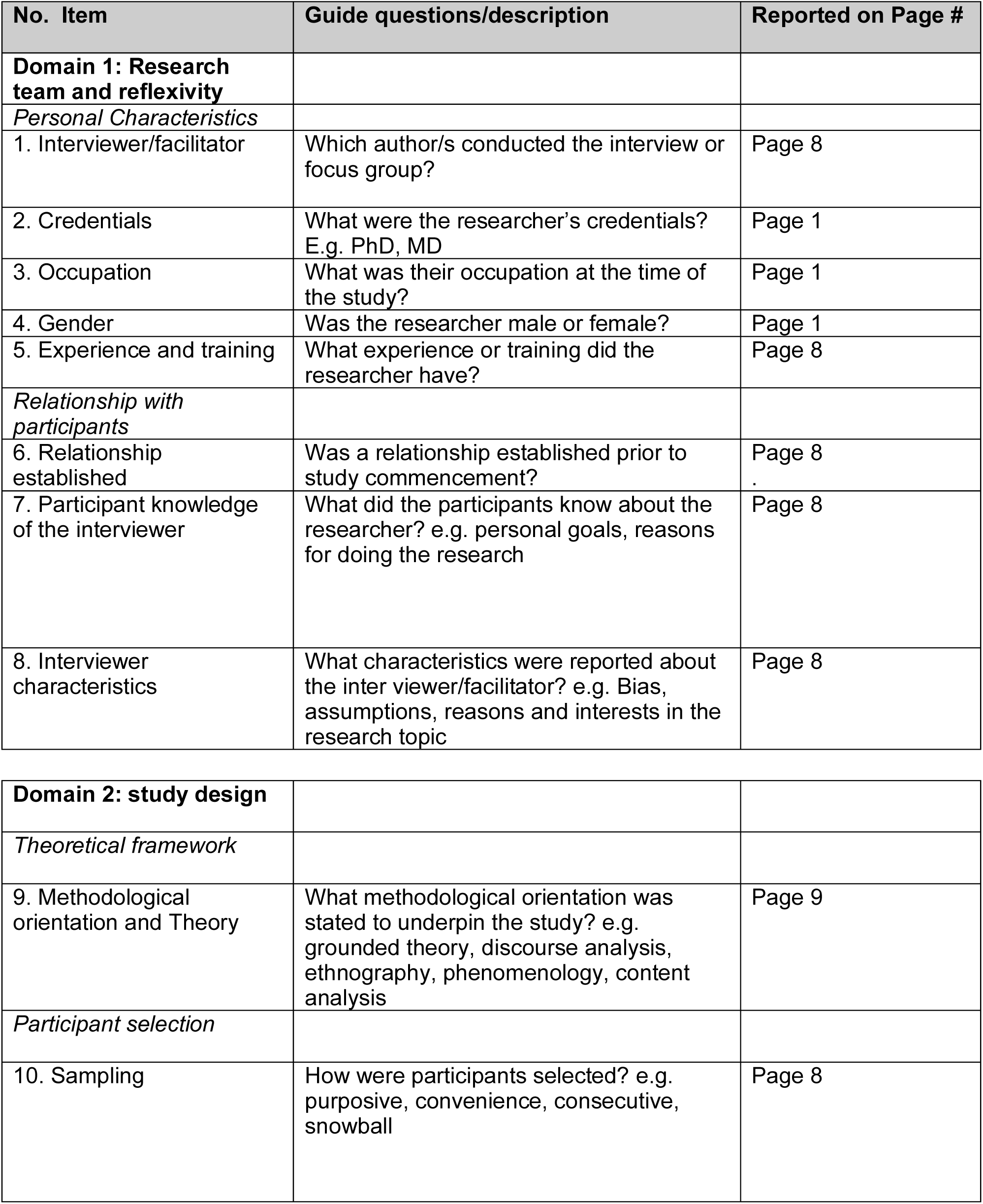
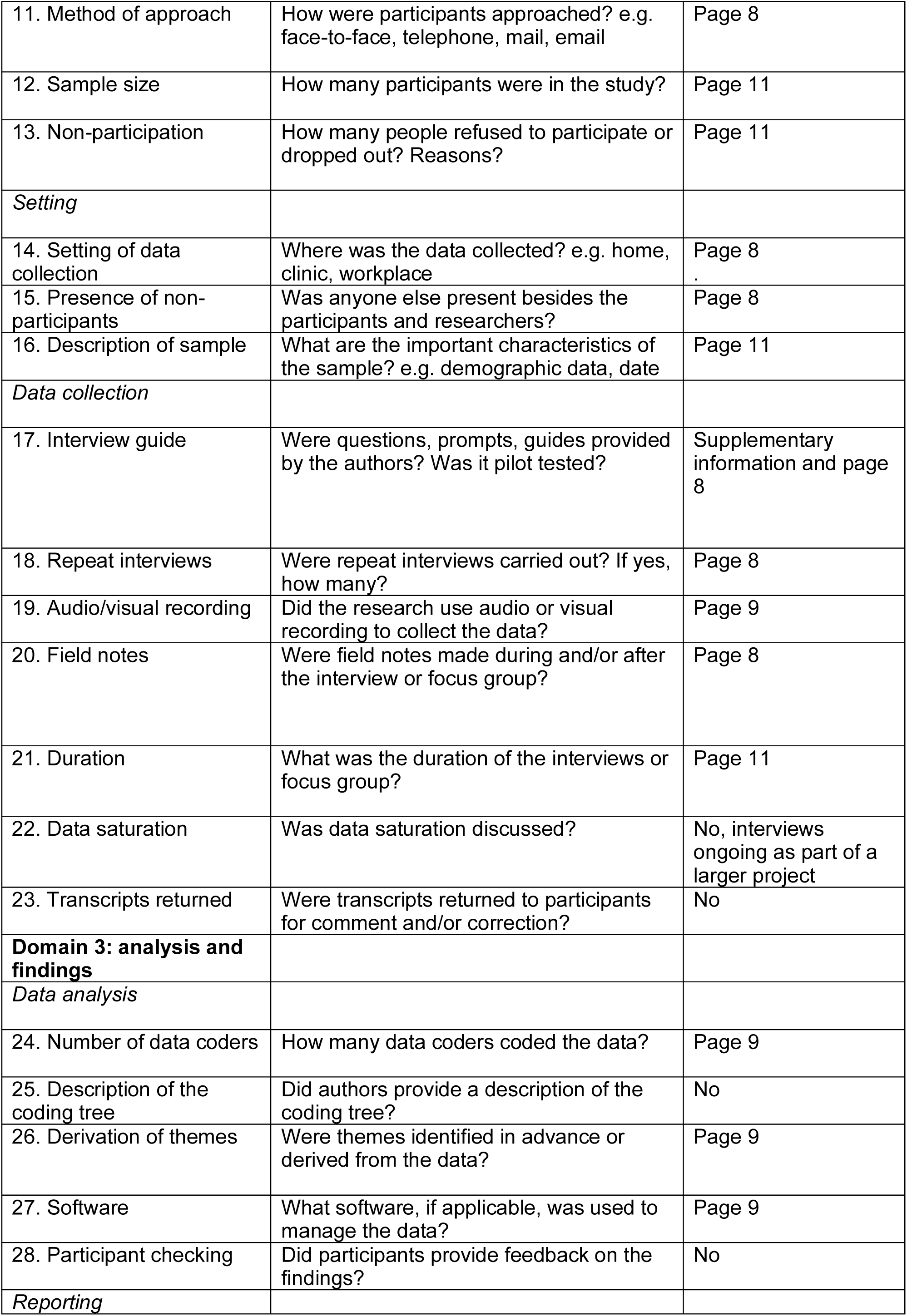
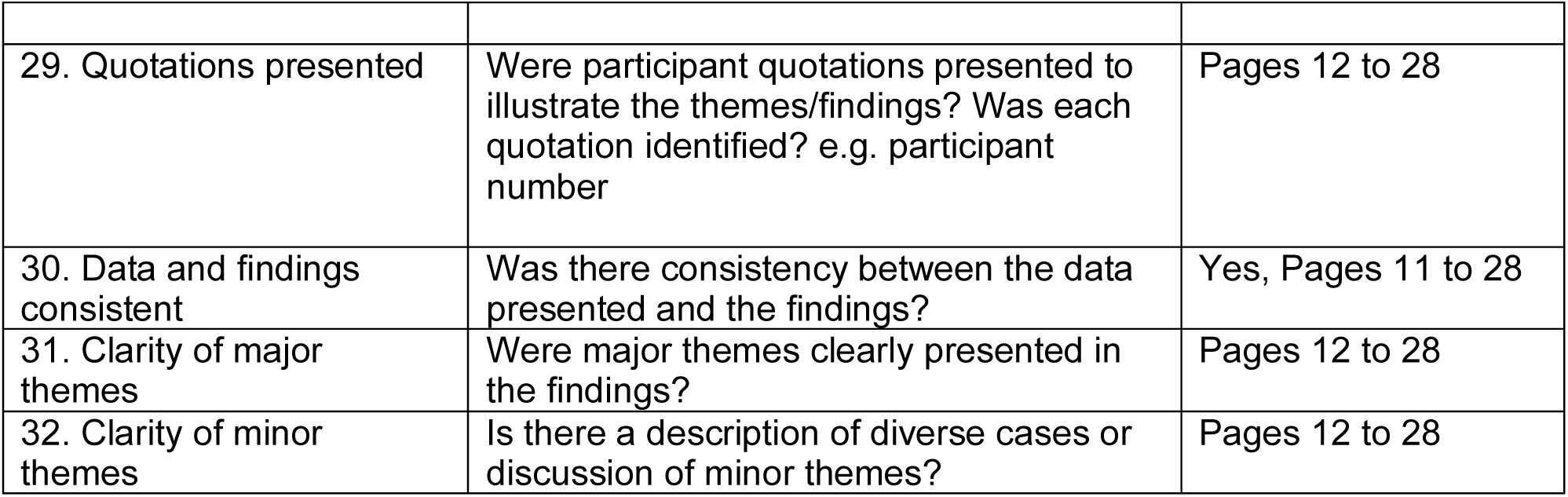

